# A Sex-specific Mendelian Randomization-Phenome-Wide Association Study of Body Mass Index

**DOI:** 10.1101/2024.09.12.24313524

**Authors:** Zhu Liduzi Jiesisibieke, Io Ieong Chan, Jack Chun Man Ng, C Mary Schooling

**Author notes:** Corresponding Author: C Mary Schooling Graduate School of Public Health and Health Policy, City University of New York, New York, United States. Grants or fellowships supporting the writing of the paper: The authors declare that there is no funding support regarding the publication of this paper. Disclosure summary: The authors have nothing to disclose. Impact statement: BMI may affect a wide range of health-related attributes and there are notable sex differences in its impact, including opposite associations for certain attributes, such as ApoB; and stronger effects in men, such as for cardiovascular diseases. Our findings underscore the need for nuanced, sex-specific policy related to BMI.

## Abstract

**Background:** Trials of incretins are making it increasingly clear that body mass index (BMI) is linked to several diseases throughout life, but trials cannot easily provide a comprehensive assessment of the role of BMI in health-related attributes for men and women. To systematically investigate the role of BMI, we conducted a sex-specific Mendelian randomization-phenome-wide association study.

**Methods:** We comprehensively examined the associations of genetically predicted BMI in women (n: 194,174) and men (n: 167,020) using health-related attributes from the UK Biobank with inverse variance weighting and sensitivity analysis.

**Results:** BMI impacted 232 of 776 traits considered in women and 203 of 680 traits in men, after adjusting for false discovery; differences by sex were found for 105 traits, and 46 traits remained after adjusting for false discovery. BMI was more strongly positively associated with myocardial infarction, major coronary heart disease events, ischemic heart disease and heart attack in men than women. BMI was more strongly positively associated with apolipoprotein B (ApoB) and diastolic blood pressure in women than men.

**Conclusion:** Our study revealed that BMI might affect a wide range of health-related attributes and also highlights notable sex differences in its impact, including opposite associations for certain attributes, such as ApoB; and stronger effects in men, such as for cardiovascular diseases. Our findings underscore the need for nuanced, sex-specific policy related to BMI to address inequities in health.

Funding: None

## Introduction

Global obesity prevalence more than doubled from 1980 to 2008 (Finucane et al., 2011). In 2022, one in eight people were obese worldwide (WHO, 2024). Body mass index (BMI) is a longstanding measure of obesity, despite its high specificity but low sensitivity (Chooi et al., 2019; Okorodudu et al., 2010). Many observational studies have found BMI associated with disease risk factors and lifespan (Collaboration, 2009; Kahn et al., 2006; Khan et al., 2018). Generally, a higher BMI is detrimental to health. However, BMI does not account well for fat distribution. For men and women with the same BMI, women tend to store more fat (Power & Schulkin, 2008). Furthermore, men tend to store fat around their organs, while women are more likely to store it subcutaneously (Power & Schulkin, 2008), potentially leading to different health impacts by sex (Costanzo et al., 2022; Power & Schulkin, 2008). As such, the effect of BMI on health may vary between men and women and may be a modifiable factor contributing to differences in lifespan by sex.

Few previous observational studies have systematically assessed the role of BMI in health and disease, and are limited by their susceptibility to confounding (Fewell et al., 2007). Mendelian randomization (MR) studies reduce confounding by using genetic proxies, such as single nucleotide polymorphisms (SNPs), for exposures (Smith & Ebrahim, 2003). Previous phenome-wide association studies using MR (MR-PheWASs) have identified impacts of sex-combined BMI on endocrine disorders, circulatory diseases, inflammatory and dermatological conditions, some biomarkers and feelings of nervousness (Hyppönen et al., 2019; Millard et al., 2019; Millard et al., 2015), but did not systematically use sex-specific BMI for the exposure or sex-specific outcomes. Previous MR studies and trials of incretins have expanded our knowledge about a broad range of effects of BMI (Larsson et al., 2020; Marso et al., 2016). To our knowledge, no sex-specific PheWAS has investigated the effects of BMI on health outcomes (Hyppönen et al., 2019; Millard et al., 2019; Millard et al., 2015). To address this gap, we conducted a sex-specific PheWAS, using the largest available sex-specific GWAS of BMI, to explore the impact of sex-specific BMI on sex-specific health-related attributes.

## Methods

### Data sources

#### Exposure: Body mass index

To obtain genetic information for BMI sex-specifically, we used BMI (inverse rank normalized) from two sources: a prospective cohort study of half a million adults from the UK Biobank, and the Genetic Investigation of ANthropometric Traits (GIANT) consortium (Locke et al., 2015), a GWAS meta-analysis of 339,224 participants mostly of European ancestry. The UK Biobank has the advantage of a larger sample size and denser genotyping but is the only large-scale sex-specific source for many outcomes. The sex-specific UK Biobank BMI GWAS conducted by Neale Lab (women: 194,174; men: 167,020) was adjusted for age, age squared and the first 20 principal components (https://www.nealelab.is/uk-biobank/faq). The GIANT GWAS has the advantage of using a different sample from the UK Biobank but is smaller with less dense genotyping (women: 171,977; men: 152,893). The sex-specific GIANT GWAS was adjusted for age, age squared, and study-specific covariates (Locke et al., 2015). We also considered overall BMI from the GIANT (n: 681,275) which includes the UK Biobank participants (approximately 64%), and was adjusted for age, age squared, principal components and study-specific covariates (Yengo et al., 2018).

### Outcomes

The UK Biobank is currently the largest and most comprehensive source for sex-specific GWAS, provided by Neale Lab (women: 194,174; men: 167,020), including many disease outcomes and physiological attributes. The average age at recruitment of UK Biobank participants was 57 years, as described previously (Collins, 2012).

### Outcomes: Inclusion and exclusion criteria

For continuous outcomes only rank normalized health attributes with at least 1,000 participants (Verma et al., 2018) were included to maintain statistical power. For binary attributes only those with at least 200 cases were included, while duplicate phenotypes were excluded. Where GWAS of very similar attributes were provided, we only included one instance. Where an attribute has a standard measure, such as forced expiratory volume in 1 second, we used that in preference to similar measures. We also excluded attributes only assessed in selected subgroups of the UK Biobank participants, such as electrocardiogram, which was only conducted in healthy people (Ramírez et al., 2021). Where attributes were available from both self-report and doctor diagnosis, we used self-reports. This is because comprehensive record linkage to doctor diagnoses has not yet been fully implemented for the UK Biobank, so information from doctor diagnoses may not fully represent the broader UK Biobank cohort. Finally, attributes with known measurement issues, such as estrogen and immature reticulocyte fraction, were excluded (Newman & Handelsman, 2014; Piva et al., 2015). Detailed exclusion criteria are listed in **Figure 1**.

**Figure 1.**
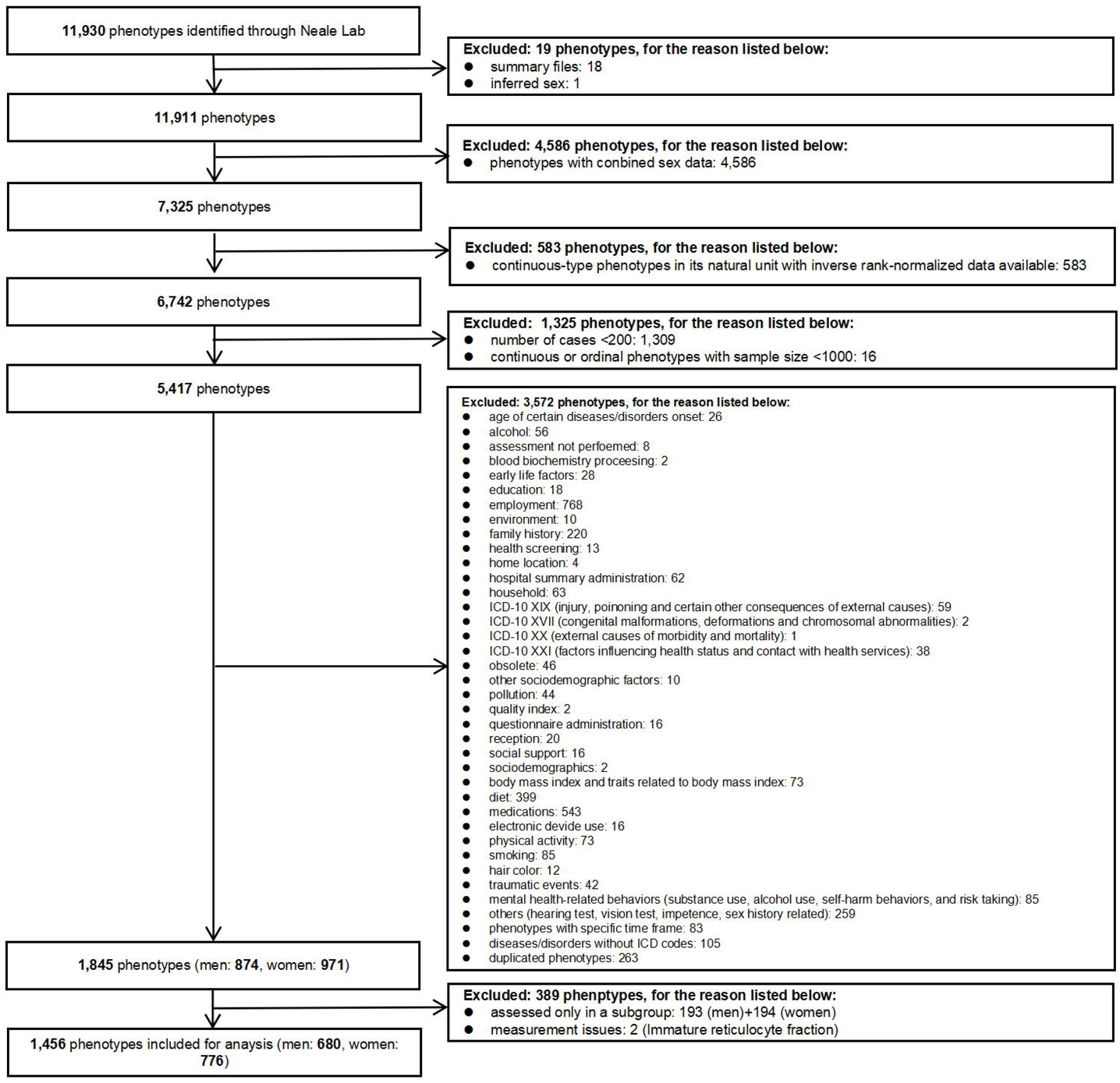
Selection criteria for the phenotypes

### Outcomes: Attribute categorization

We categorized attributes as age at recruitment, physical measures, lifestyle and environmental, medical conditions, operations, physiological factors, cognitive function, health and medical history, sex-specific factors, blood assays and urine assays based on the UK Biobank categories (https://biobank.ndph.ox.ac.uk/ukb/cats.cgi). Binary attributes with an International Classification of Diseases-10 code were categorized by International Classification of Diseases-10 chapter, as (I) infectious diseases, (II) neoplasms, (III) diseases of the hematopoietic system and blood disorders, (IV) endocrine and metabolic diseases, (V) psychological disorders, (VI) diseases of the nervous system, (VII and VIII) diseases of the sensory system (eyes and ears), (IX) diseases of the circulatory system, (X) diseases of the respiratory system, (XI) diseases of the digestive system, (XII) diseases of skin and subcutaneous tissue, (XIII) diseases of the musculoskeletal system and connective tissue, (XIV) diseases of the genitourinary system, (XV) pregnancy, childbirth, and the puerperium and (XVIII) symptoms.

#### Statistical analysis

MR was used to assess effects of genetically predicted sex-specific BMI on each attribute considered sex-specifically. Inverse-variance weighted estimates were used initially, i.e., meta-analysis of Wald estimates (SNP on outcome divided by SNP on exposure), and then sensitivity analysis (MR-Egger) was used for any associations found. The significance level was adjusted for the false discovery to account for multiple comparisons (Benjamini & Hochberg, 1995). Specifically, we ranked the p-values for men and women respectively, calculated the Benjamini-Hochberg (BH) value, and identified the significant attributes influenced by BMI using the BH value. We obtained differences by sex using a z-test (Paternoster et al., 1998), which as recommended was on a linear scale for dichotomous outcomes (Knol et al., 2007; Rothman, 2008), then we identified which ones remained after allowing for false discovery. We used the R packages “TwoSampleMR” (version: 0.6.1), “Mendelian Randomization” (version: 0.10.0) for the MR analysis and “metafor” (version: 4.6-0) to assess sex-differences.

## Results

Initial analysis using sex-specific BMI from GIANT yielded similar estimates as when using sex-specific BMI from the UK Biobank but had fewer SNPs resulting in wider confidence intervals (**S Table 1**) and fewer significant associations (**S Table 2**). Analysis using sex-combined GIANT yielded more significant associations but lacks granularity, so we presented the results obtained using sex-specific BMI from the UK Biobank.

### Sex-specific estimates

In men, BMI was associated with 203 of the 680 health-related attributes considered using false discovery (Figure 2, **S Table 2**). As expected, BMI was positively associated with ischemic heart disease, heart failure, hypertensive heart and/or renal disease, major coronary heart disease events, heart attack, diabetes, hypertension, sleep apnoea, daytime dozing/sleeping (narcolepsy), triglycerides, urinary potassium, urinary sodium and major surgeries. BMI was also inversely associated with age at recruitment, osteoporosis, hay fever and allergic rhinitis or eczema, high density lipoprotein cholesterol (HDL-c), cholesterol, low density lipoprotein cholesterol (LDL-c), sex hormone binding globulin (SHBG) and total testosterone. Positive associations of BMI with higher risk of digestive system cancers, lymphomas, primary lymphoid and hematopoietic malignancies, and esophageal cancer were also found in men. Furthermore, we found higher BMI associated with accelerated facial aging and baldness.

**Figure 2.**
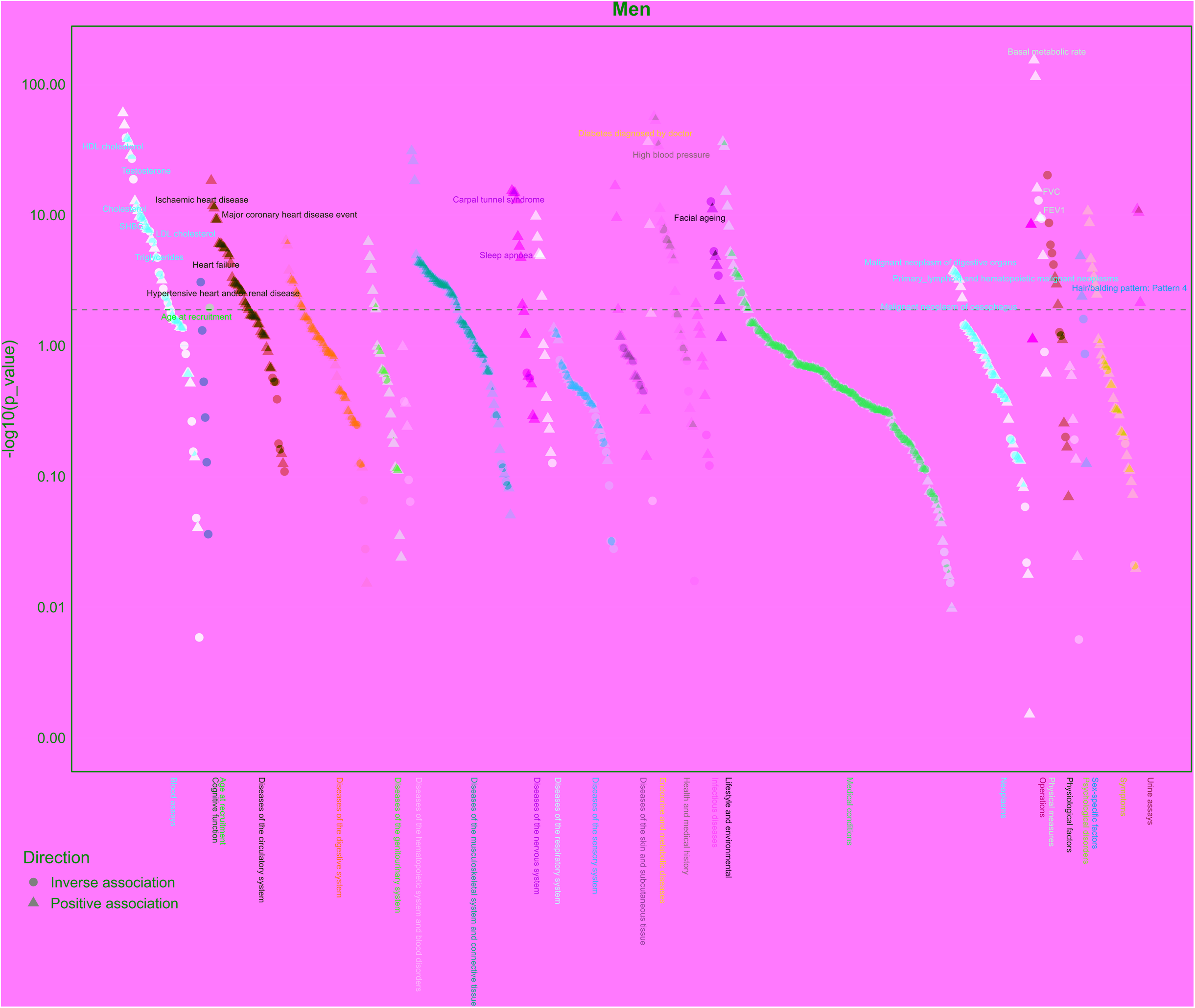
Manhattan plot of body mass index on phenotypes in men (n: 167,020) from the UK Biobank. Note: The dotted lines indicate the significant threshold.

In women, BMI was associated with 232 of 776 health-related attributes considered using the false discovery (Figure 3, **S Table 2)**. As expected, BMI was positively associated with ischemic heart disease, heart failure, hypertensive heart and/or renal disease, heart attack, diabetes, hypertension, sleep apnoea, daytime dozing/sleeping (narcolepsy), apolipoprotein B (ApoB), triglycerides, total testosterone and urinary potassium and sodium and major surgeries. BMI was also inversely associated with osteoporosis, HDL-c, cholesterol, LDL-c and SHBG. BMI was positively associated with risk of uterine/endometria cancer, cancer of the bronchus and lung and intrathoracic organs, as well as cancers affecting the skin and female reproductive organs. Higher BMI in women was also associated with accelerated facial aging.

**Figure 3.**
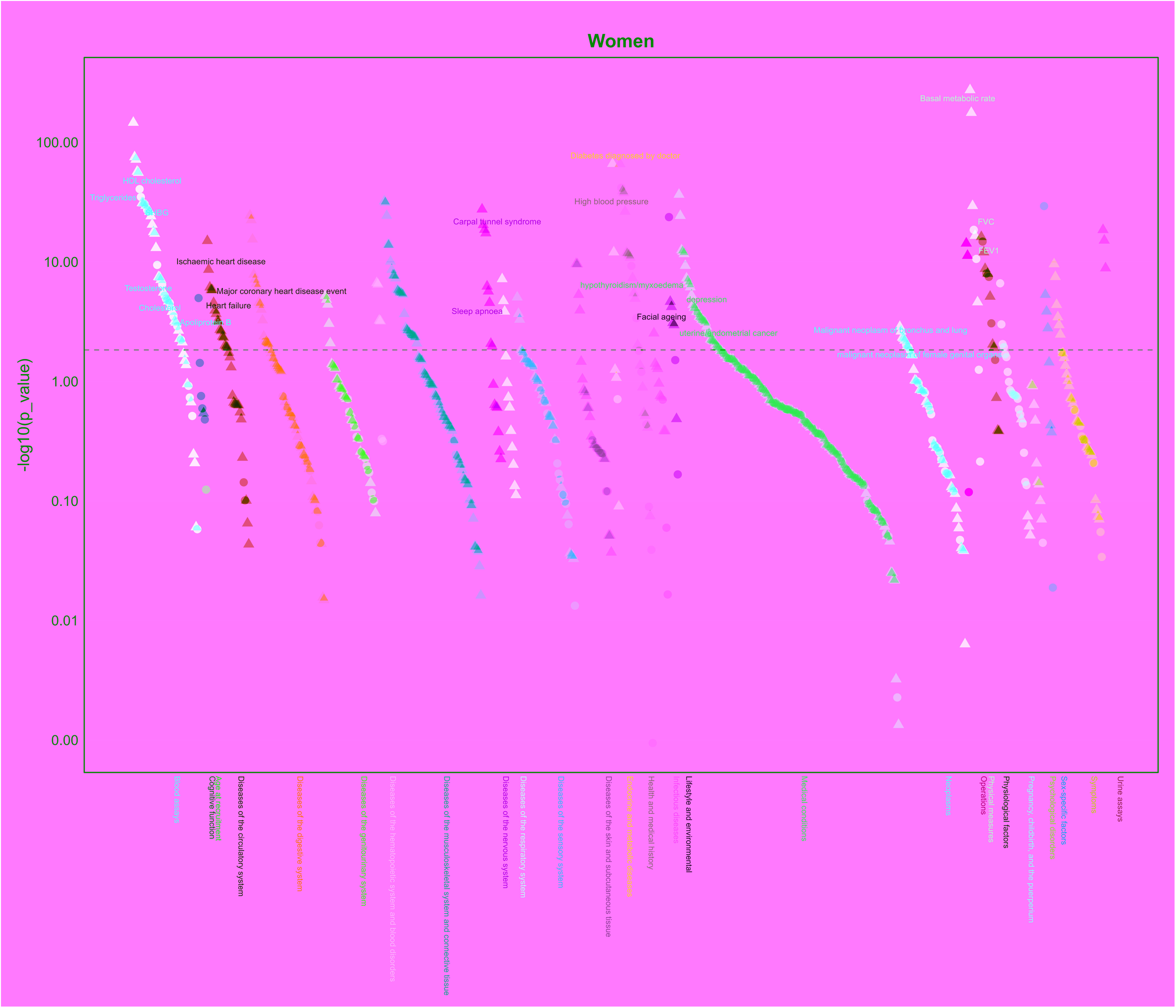
Manhattan plot of body mass index on phenotypes in women (n: 194,174) from the UK Biobank. Note: The dotted lines indicate the significant threshold.

### Differences by sex

We found significant differences by sex in the associations of BMI with 105 health-related attributes (p-value<0.05); 46 phenotypes remained after allowing for false discovery (**Table 1**). Of these 46 differences most (35) were in magnitude but not direction, such as for SHBG, ischemic heart disease, heart attack, and facial aging, while 11 were directionally different.

**Table 1.**
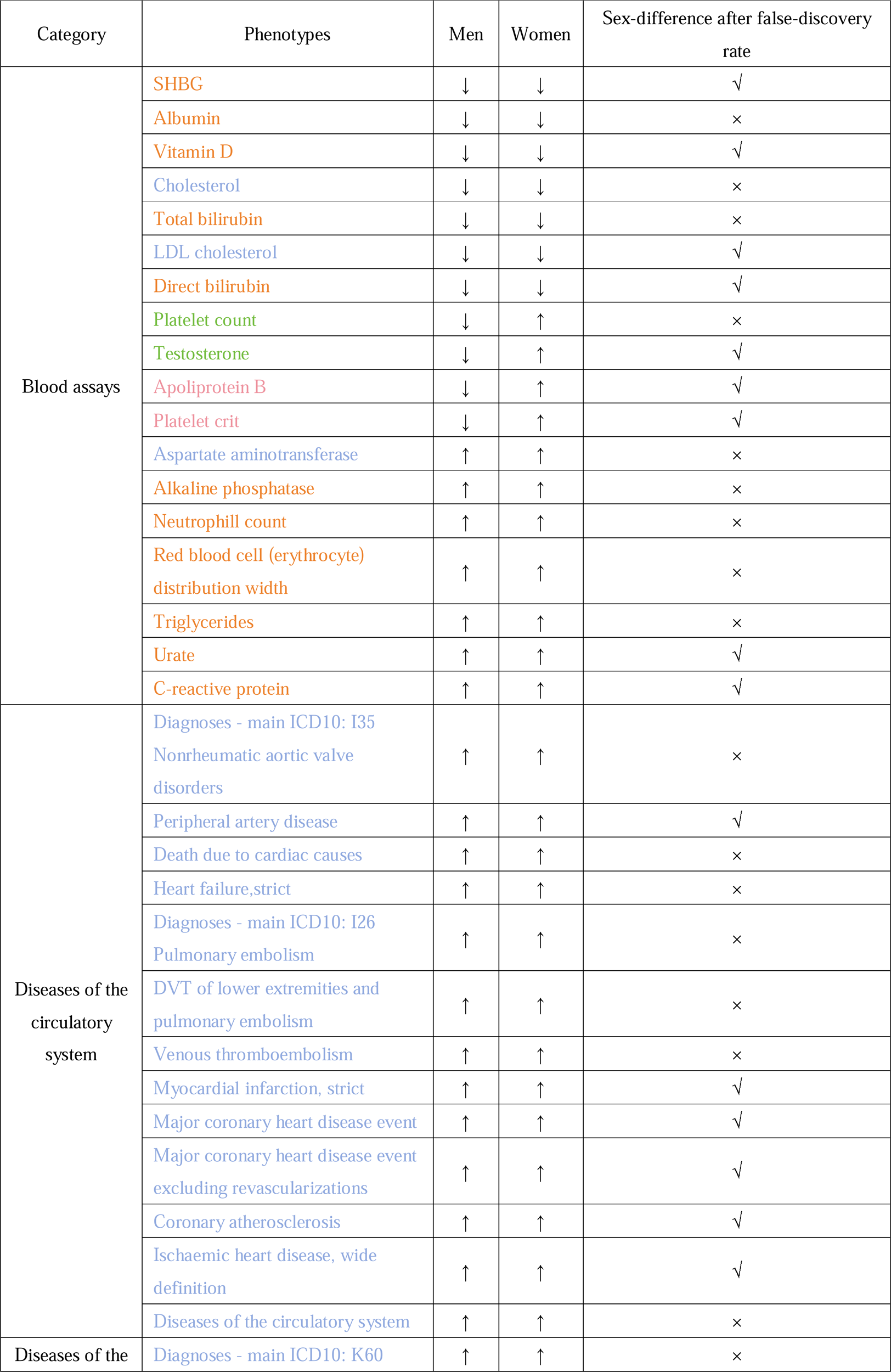
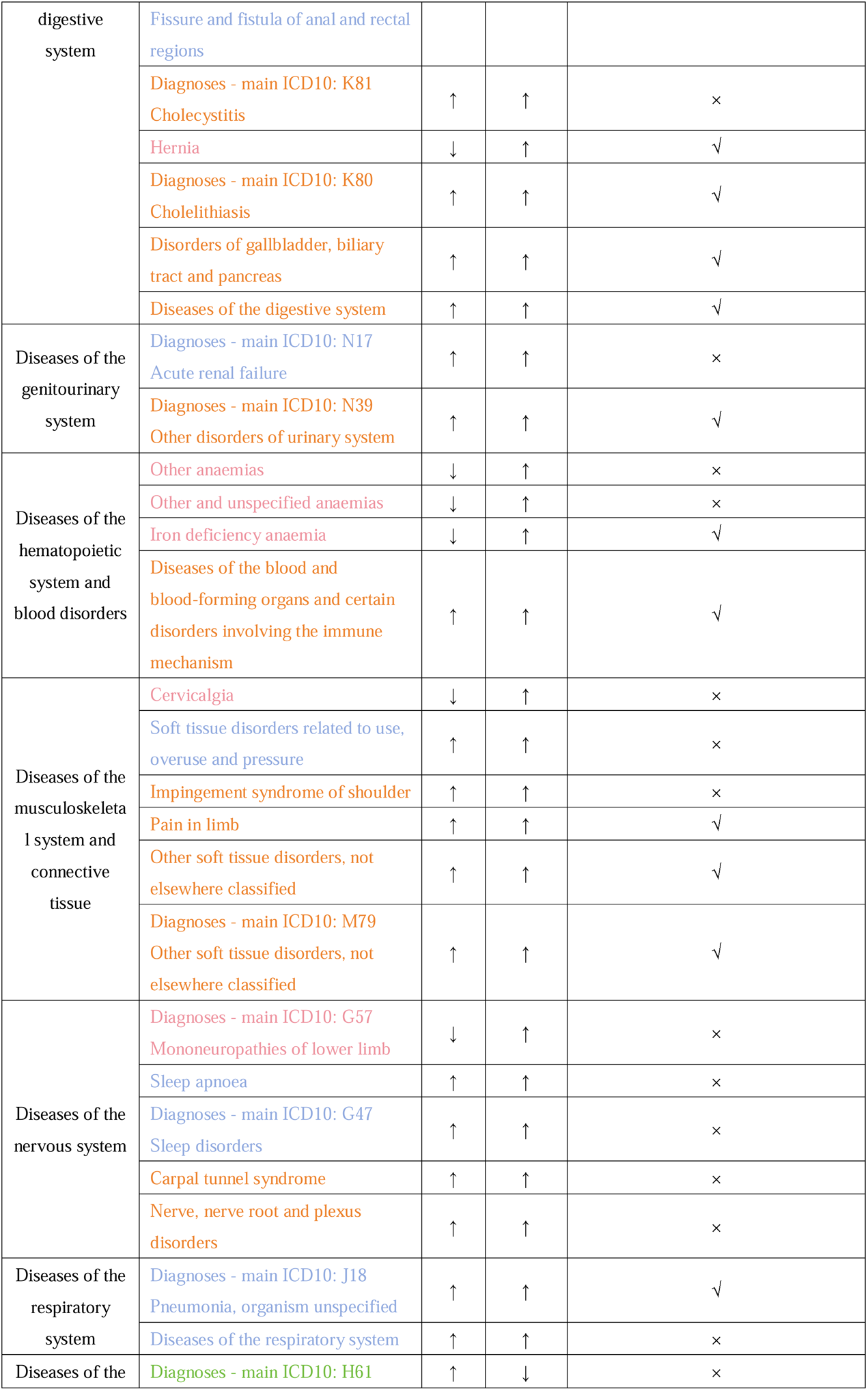
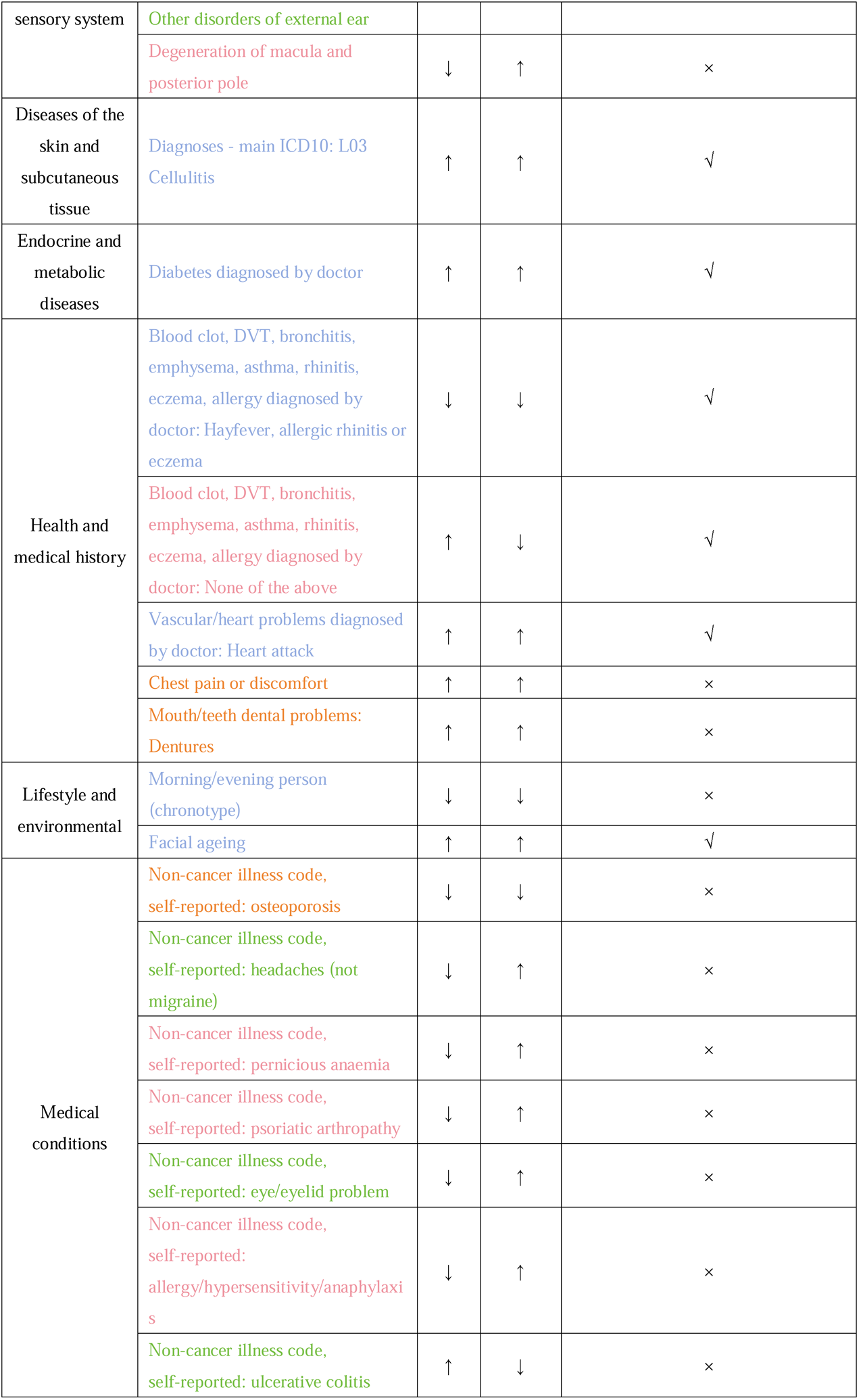
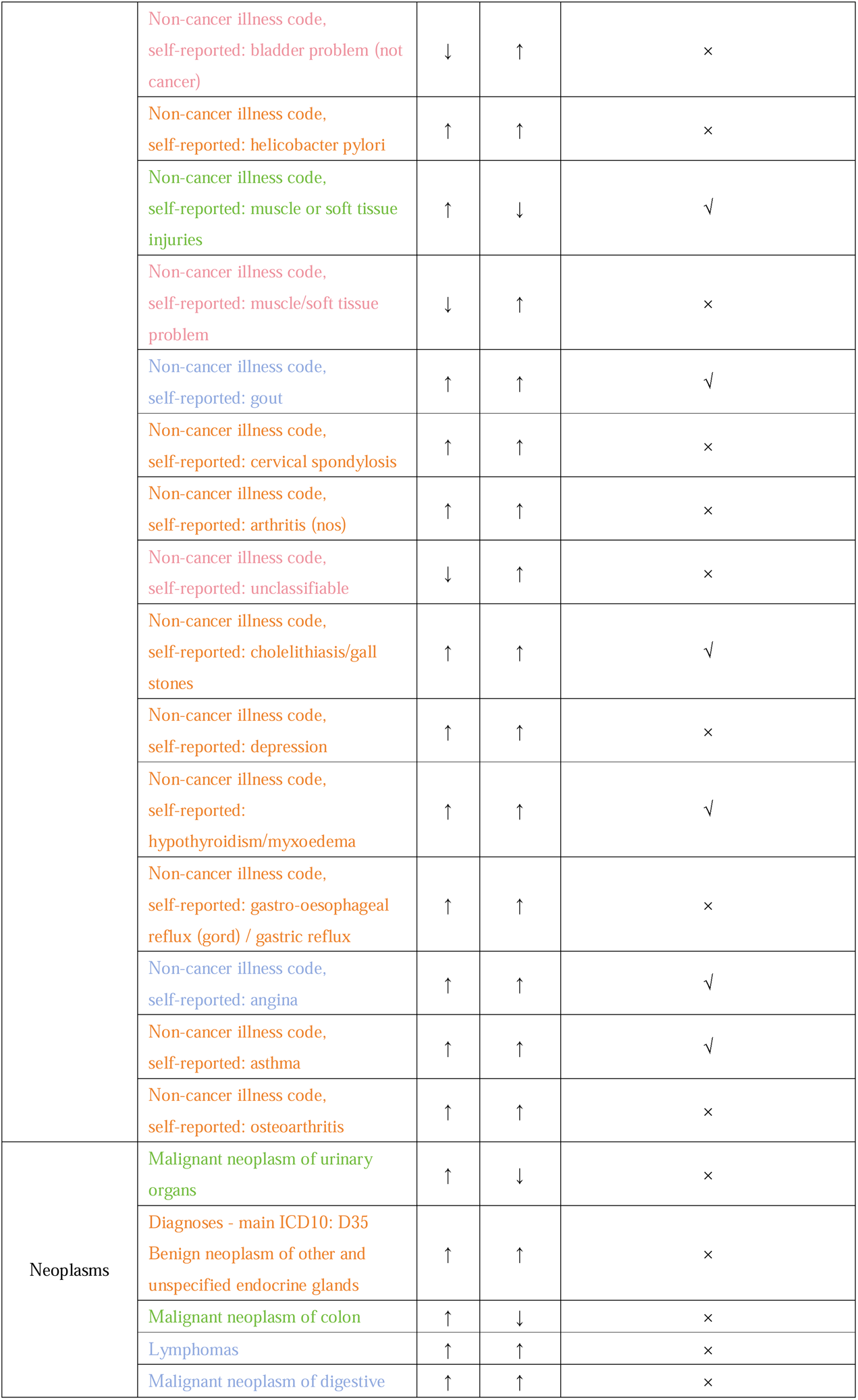
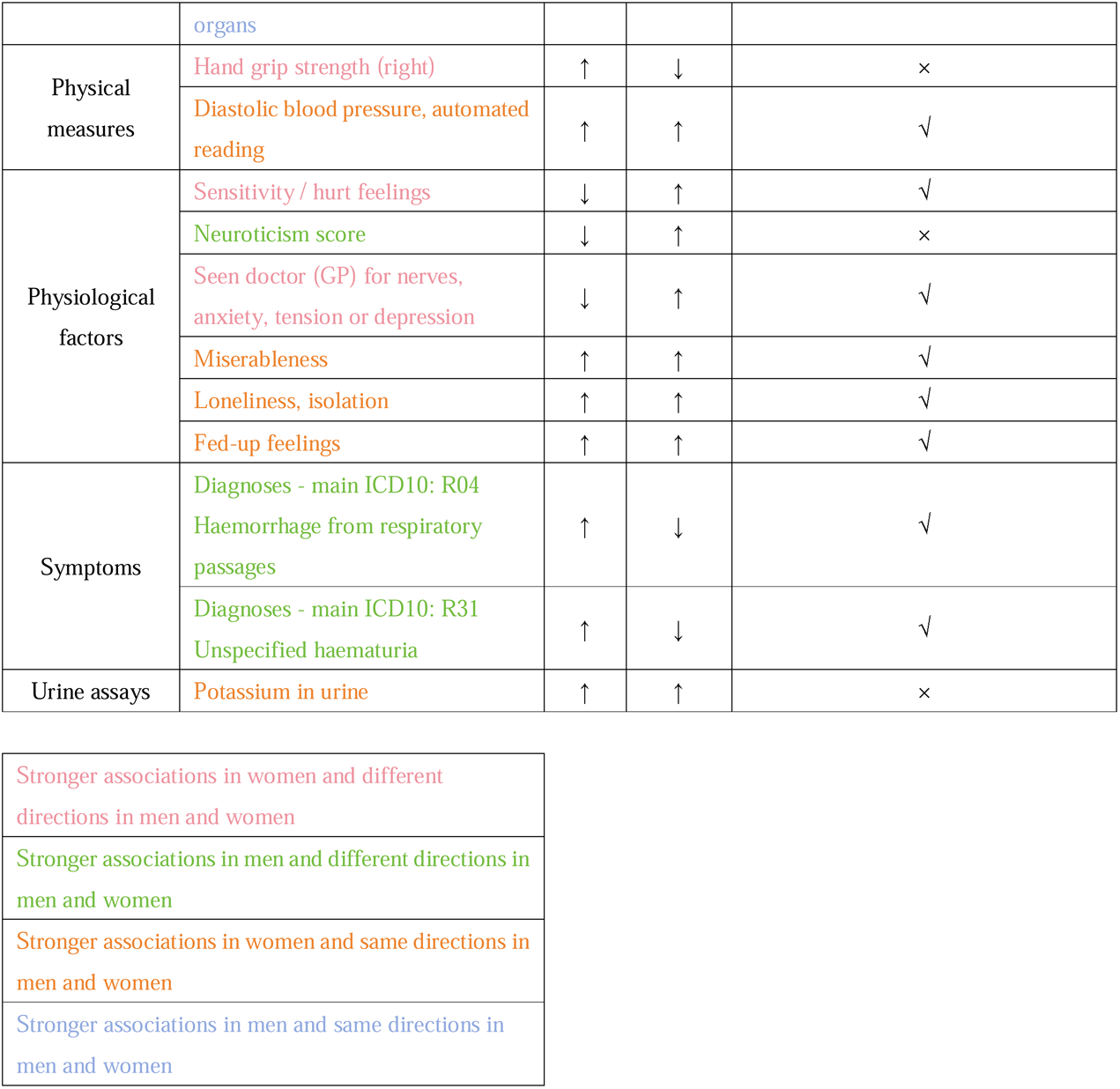
Sex differences of body mass index on phenotypes in men (n: 167,020) and women (n: 194,174)

Notably, BMI was more strongly positively associated with myocardial infarction, major coronary heart disease events, ischemic heart disease, heart attack, and facial aging in men than in women. BMI was more strongly positively associated with diastolic blood pressure, and hypothyroidism/myxoedema in women than men. BMI was more strongly inversely associated with LDL-c, hay fever and allergic rhinitis or eczema in men than women. BMI was more strongly inversely associated with SHBG in women than men.

BMI was inversely associated with ApoB, iron deficiency anemia, hernia, and total testosterone in men, while positively associated with these traits in women (**Table 1**). BMI was inversely associated with sensitivity/hurt feelings, and ever seeking medical advice for nerves, anxiety, tension, or depression in men. However, BMI was positively associated with sensitivity/hurt feelings and ever seeking medical advice for these same issues in women. BMI was positively associated with muscle or soft tissue injuries and haemorrhage from respiratory passages in men, whilst inversely associated with these traits in women.

Of the 42 significant sex-specific associations identified in both the UK Biobank and the sex-specific GIANT consortium for men, all were directionally consistent. Similarly, for women, all 45 such significant associations were directionally consistent.

## Discussion

Consistent with previous studies BMI was positively associated with many health-related attributes, such as ischemic heart disease, heart failure, hypertensive heart and/or renal disease, diabetes, hypertension and sleep apnoea. Our study adds by showing sex-differences in some traits related to psychological disorders as well as ApoB and showing stronger associations in men than women for common cardiovascular diseases.

### Comparison with previous studies

Consistent with previous MR studies, BMI was positively associated with the risk of ischemic heart disease, heart failure, hypertensive heart disease (Riaz et al., 2018), hypertension (Riaz et al., 2018) and diabetes (Larsson & Burgess, 2021) in both men and women as would be expected. A previous MR study found height and fat-free mass positively associated with follicular lymphoma (Zhou et al., 2024). However, we found BMI positively associated with lymphomas in men, but not in women (although directionally consistent and not significant after allowing for false discovery). Another MR study found BMI positively associated with asthma and poorer lung function, but not with hay fever (Skaaby et al., 2018), while we found BMI inversely associated with hay fever and allergy allergic rhinitis or eczema in men, but not in women. Previous studies have shown BMI inversely associated with sodium/potassium (Zanetti et al., 2020), while we found BMI positively associated with urinary potassium and sodium. Additionally, we found a positive association of BMI with major surgeries. Consistent with a previous MR study (Ardissino et al., 2022), we found BMI positively associated with sleep apnoea. We also found positive associations of BMI with daytime dozing in both sexes, which may indicate effects of BMI on quality of life. Higher BMI was also associated with a younger age at recruitment among men, suggesting poorer survival to recruitment in men than women given the GWAS adjusted for age and the short recruitment window makes period effects unlikely. The association of BMI with age at recruitment was less evident in women, with no significant sex difference. Consistently, a prior observational study found high BMI more detrimental to lifespan in men than women, especially at younger ages (Fontaine et al., 2003).

### Differences in the associations of body mass index with health-related attributes in men and women

BMI was more strongly associated with common cardiovascular diseases in men than women. BMI was positively associated with ApoB in women, whereas in men, the association was inverse (but not significant after correction for multiple comparisons). However, the sex difference was significant. This pattern contrasts with earlier MR studies that reported an inverse association of BMI with ApoB in the general population (Bell et al., 2022). BMI being more strongly associated with heart disease in men than women while also being protective for ApoB, a key heart disease risk factor, requires some explanation. BMI is a complex phenotype that may represent different attributes or have different consequences in men compared with women. Fat mass storage tends to differ between men and women (visceral versus subcutaneous) (Power & Schulkin, 2008). The causes of higher BMI may also differ between men and women, due to occupational roles, gender-based food preferences or cultural norms (Kanter & Caballero, 2012). Alternatively, unknown factors affected by BMI could contribute to heart disease specifically to men. Whether the difference in ischemic heart disease rates between men and women that emerged in the US and the UK the late 19^th^ century (Nikiforov & Mamaev, 1998) is explained by rising BMI remains to be determined. In terms of psychological disorders, women being more affected psychologically by higher BMI is consistent with sociocultural pressures and norms surrounding women’s rather than men’s bodies (Esnaola et al., 2010; Schwartz & Brownell, 2004). We also found that BMI was associated with balding in men but not women, although the sex difference was not significant Moreover, BMI was positively associated with facial aging in both men and women, with larger effects in men.

### Strengths and limitations

A major strength of this study was the focus on sex differences, which have not been explicitly explored in previous PheWAS of BMI, despite differences in lifespan by sex. We also considered false discovery for sex differences. Despite the large sample size and the broad range of outcomes considered in this study, limitations and concerns exist. First, MR has stringent assumptions, i.e., relevance, independence and exclusion restriction. The F-statistics were above 10, which addresses relevance. MR is largely free from confounding by design. For a few attributes (37 out of 1,456), the MR-Egger intercept was significant while the IVW estimate was not (**S Table 3**), potentially indicating horizontal pleiotropic effects or the play of chance, which requires further investigation. Second, MR assesses lifetime effects of BMI, which may differ from those of short-term weight changes or weight cycling. Third, not all attributes were included. For example, heart rate from electrocardiogram (ECG) and age at asthma diagnosis were excluded, because such information was limited to selected subgroups. Fourth, given the study’s exploratory nature, we allowed for multiple comparisons to minimize chance findings, which means some BMI effects may not be captured. However, such comprehensive consideration may identify previously unknown or ignored impacts, such as facial aging and balding. Fifth, while the UK Biobank does not fully represent the UK population, representativeness is not crucial for causal inference as long as the sample is not selected on exposure and outcome (Greenland, 2003). Sixth, focusing on a European population may limit generalizability, but it helps reduce bias from population stratification. Seventh, we excluded replicated or very similar attributes, potentially missing some attributes. However, this reduces false negatives when adjusting for multiple comparisons. Eighth, this study is open to selection bias because genetic endowment is lifelong but the UK Biobank participants are middle-aged or older, so potential recruits who did not live to recruitment because of their BMI are missing. This may bias estimates towards the null or inverse for harmful binary outcomes (Thompson et al., 2013), but is less likely to affect continuous outcomes (Smit et al., 2019). Ninth, we focused on BMI, because it is well accepted and easy to measure, although waist-to-hip ratio may be a better marker for mortality (Khan et al., 2023; Ruder, 2023). Tenth, although this study primarily utilized sex-specific BMI, we also conducted analyses using overall BMI from GIANT including the UK Biobank, which gave a generally similar interpretation (**S Table 1**). Using sex-specific BMI from the UK Biobank and GIANT may lead to lower statistical power than using overall population BMI but allows for the detection of traits that are affected differently by BMI by sex. Including findings from the overall population BMI from sex-combined GIANT (**S Table 1**) makes the results more comparable to previous similar studies. Eleventh, our study did not find associations of BMI with some early-onset cancers possibly because early-onset cancers tend to be of germline origin (Qing et al., 2020). Twelfth, we used information from Neale Lab, whose quality check removed participants whose self-reported sex differed from their biological sex, so our findings only relate to *cis* men and women. Thirteenth, while overlapping samples for BMI and the outcomes pose a potential issue in the main analysis, we also replicated the analysis using sex-specific instruments for BMI independent of the UK Biobank. Results were similar, suggesting bias from overlapping samples is minimal. Consistently, overlapping samples have been shown to make most difference for relatively small samples and for MR-Egger estimates, particularly when the I^2^_GX_ is low, leading to confounded estimates (Minelli et al., 2021). Lastly, we used linear MR, so we cannot exclude the possibility of a “J” shaped association, however trials of incretins suggest a “J” curve may be a manifestation of bias (Wilding et al., 2021).

### Public health implications

Trials of incretins have clearly indicated many benefits of weight loss particularly for older people (Leiter et al., 2019; Lincoff et al., 2023). Our study showing how BMI might increase risk of chronic diseases, reduce quality of life, and depress mental health in women, underscores the importance of addressing the obesity epidemic equitably in men and women, given differences by sex in some consequences of overweight/obesity, such as psychological disorders and lipids. Being overweight or obese is more common in men than women in developed countries (Kanter & Caballero, 2012; Maruyama & Nakamura, 2018). With ongoing global economic development, the same pattern is likely to become more common. Currently women are more likely to seek medical intervention for weight management, such as semaglutide, than men (Luthra, 2023). To promote population health, it is important to address the unique challenges faced by overweight men and women. Men are at higher risk of heart disease and premature death due to high BMI, so targeted public health interventions, such as sex-specific weight recommendations or guidelines, could perhaps be considered.

## Conclusion

Our contemporary systematic examination found BMI associated with a broad range of health-related attributes. We also found significant sex differences in many traits, such as for cardiovascular diseases, underscoring the importance of addressing higher BMI in both men and women possibly as means of redressing differences in life expectancy. Ultimately, our study emphasizes the harmful effects of obesity and the importance of nuanced, sex-specific policy related to BMI to address inequities.in health.

## Data Availability

This study used data from the MR-base platform (https://www.mrbase.org/), UK Biobank (http://www.nealelab.is/uk-biobank/).

## Abbreviations

ApoB: apolipoprotein B
BMI: body mass index
ECG: electrocardiogram (ECG)
GIANT: Genetic Investigation of ANthropometric Traits
GWAS: genome-wide association study
HDL-c: high density lipoprotein cholesterol
LDL-c: low density lipoprotein cholesterol
MR: Mendelian Randomization
SHBG: sex hormone binding globulin

## Acknowledgements

We would like to thank the Neale Lab for providing GWAS summary-level statistics.

## Consent for publication

All participants consented to publication in accordance with the original datasets (publicly available databases).

## Ethical approval

The study protocol was not pre-registered. We used only publicly available summary-level data and did not collect any original data in this study. Ethics approval and consent from individual participants can be found in the original publications.

## Credit author statement

Conceptualization: Zhu Liduzi Jiesisibieke, C Mary Schooling

Data curation: Zhu Liduzi Jiesisibieke, C Mary Schooling, Io Ieong Chan, Jack Chun Man Ng

Formal analysis: Zhu Liduzi Jiesisibieke, C Mary Schooling

Methodology: Zhu Liduzi Jiesisibieke, C Mary Schooling, Io Ieong Chan, Jack Chun Man Ng

Project administration: C Mary Schooling Resources: C Mary Schooling

Software: Zhu Liduzi Jiesisibieke, C Mary Schooling Supervision: C Mary Schooling

Visualization: Zhu Liduzi Jiesisibieke, C Mary Schooling

Writing – original draft: Zhu Liduzi Jiesisibieke, C Mary Schooling Writing – review & editing: Zhu Liduzi Jiesisibieke, C Mary Schooling

